# COVID-19 rebound after Paxlovid and Molnupiravir during January-June 2022

**DOI:** 10.1101/2022.06.21.22276724

**Authors:** Lindsey Wang, Nathan A. Berger, Pamela B. Davis, David C. Kaelber, Nora D. Volkow, Rong Xu

## Abstract

**Importance:** Recent case reports document that some patients who were treated with Paxlovid experienced rebound COVID-19 infections and symptoms 2 to 8 days after completing a 5-day course of Paxlovid. The Centers for Disease Control and Prevention (CDC) has recently issued a Health Alert Network Health Advisory to update the public on the potential for COVID-19 rebound after Paxlovid treatments. However, the rates of COVID-19 rebound in a real-world population or whether rebound is unique to Paxlovid remains unknown.

**Objectives:** To examine the rates and relative risks of COVID-19 rebound in patients treated with Paxlovid or with Molnupiravir and to compare characteristics of patients who experienced COVID-19 rebound to those who did not.

**Design, Setting, and Participants:** Retrospective cohort study of electronic health records (EHRs) of 92 million patients from a multicenter and nationwide database in the US. The study population comprised 13,644 patients age ≥ 18 years who contracted COVID-19 between 1/1/2022-6/8/2022 and were treated with Paxlovid (n =11,270) or with Molnupiravir (n =2,374) within 5 days of their COVID-19 infection.

**Exposures:** Paxlovid or Molnupiravir.

**Main Outcomes and Measures:** Three types of COVID-19 rebound outcomes (COVID-19 infections, COVID-19 related symptoms, and hospitalizations) were examined. Hazard ratios and 95% confidence interval (CI) of 7-day and 30-day risk for COVID-19 rebound between patients treated with Paxlovid and patients treated with Molnupiravir were calculated before and after propensity-score matching.

**Results:** The 7-day and 30-day COVID-19 rebound rates after Paxlovid treatment were 3.53% and 5.40% for COVID-19 infection, 2.31% and 5.87% for COVID-19 symptoms, and 0.44% and 0.77% for hospitalizations. The 7-day and 30-day COVID-19 rebound rates after Molnupiravir treatment were 5.86% and 8.59% for COVID-19 infection, 3.75% and 8.21% for COVID-19 symptoms, and 0.84% and 1.39% for hospitalizations. After propensity-score matching, there were no significant differences in COVID-19 rebound risks between Paxlovid and Molnupiravir: infection (HR 0.90, 95% CI: 0.73-1.11), COVID-19 symptoms (HR: 1.03, 95% CI: 0.83-1.27), or hospitalizations (HR: 0.92, 95% CI: 0.56-1.55). Patients with COVID-19 rebound had significantly higher prevalence of underlying medical conditions than those without.

**Conclusions and Relevance:** COVID-19 rebound occurred both after Paxlovid and Molnupiravir, especially in patients with underlying medical conditions. This indicates that COVID-19 rebound is not unique to Paxlovid and the risks were similar for Paxlovid and Molnupiravir. For both drugs the rates of COVID-19 rebound increased with time after treatments. Our results call for continuous surveillance of COVID-19 rebound after Paxlovid and Molnupiravir treatments. Studies are necessary to determine the mechanisms underlying COVID-19 rebounds and to test dosing and duration regimes that might prevent such rebounds in vulnerable patients.

## Introduction

Paxlovid and Molnupiravir were authorized by FDA in December 2021 to treat mild-to-moderate COVID-19 in patients who are at high risk for progression to severe COVID-19^1,2^. Recently case reports have documented that some patients treated with Paxlovid experienced rebound COVID-19 infections and symptoms 2 to 8 days after completing a 5-day course of Paxlovid^3^. The Centers for Disease Control and Prevention (CDC) has issued Health Alert Network Health Advisory to update the public on the potential for COVID-19 rebound after Paxlovid treatment^4^. However, the rate of COVID-19 rebound after Paxlovid treatment in real-world population remains unknown. In addition, questions remain as to whether COVID-19 rebound is unique to Paxlovid and whether there are patients who are more susceptible. Based on a nation-wide cohort of patients who contracted COVID-19 and received Paxlovid or Molnupiravir treatment within 5 days of COVID-19 diagnosis, we examined the rates of COVID-19 rebound in patients who were treated with Paxlovid or with Molnupiravir, compared the risks for COVID-19 rebound after Paxlovid vs Molnupiravir in propensity-score matched patients, and compared characteristics of patients who experienced COVID-19 rebound to those who did not.

## Methods

### Database description

We used the TriNetX Analytics network platform that contains nation-wide and real-time de-identified electronic health records (EHRs) of 93 million unique patients from 67 health care organizations, mostly large academic medical institutions with both inpatient and outpatient facilities at multiple locations across 50 states in the US,^5^ covering diverse geographic locations, age groups, racial and ethnic groups, income levels and insurance types. TriNetX Analytics Platform performs statistical analyses on patient-level data and only reports on population level data and results without including protected health information (PHI) identifiers. MetroHealth System, Cleveland OH, Institutional Review Board has determined that research using TriNetX is not Human Subject Research and therefore exempt from review.

### Study population

The study population comprised 13,644 patients aged ≥ 18 years old who contracted COVID-19 between 1/1/2022-6/8/2022 (Omicron predominant period) who were treated Paxlovid (“Paxlovid cohort”, n =11,270) or with Molnupiravir (“Molnupiravir cohort”, n=2,374) within 5 days of their COVID-19 infection. Patients who took both drugs were excluded. The status of COVID-19 infection was based on lab-test confirmed presence of “SARS coronavirus 2 and related RNA” (Logical Observation Identifiers Names and Codes or LOINC code TNX:LAB:9088) or the International Classification of Diseases, tenth revision(ICD-10) diagnosis code of “COVID-19” (U07.1). Three COVID-19 rebound outcomes that occurred 2 days after the last day of Paxlovid or Molnupiravir were examined: (a) COVID-19 infections as determined by codes TNX:LAB:9088 or U07.1; (b) COVID-19 related symptoms^6^ including fever (ICD-10 code R50), chills (R68.83), cough (R05), shortness of breath (R06.02), fatigue (R53), muscle aches (M79.1), headache (R51), loss of taste or smell (R43), sore throat (J02), nasal congestion or rhinorrhea (R09.81), vomiting (R11.1), diarrhea (R19.7), and skin rashes (R21); (c) hospitalizations (Current Procedural Terminology or CPT code 1013659)

### Statistical analysis

The covariates are listed in **Table 1**. These covariates included demographics (age, gender, race/ethnicity); adverse socioeconomic determinants of health including problems with education, employment, occupational exposure, physical, social and psychosocial environment, and housing; medical conditions related to COVID-19 infections and adverse outcomes^7^ including comorbidities, immunosuppressant use, transplants, tobacco smoking; COVID-19 vaccination status as documented in electronic health records.

**Table 1.**
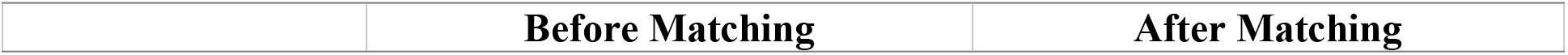
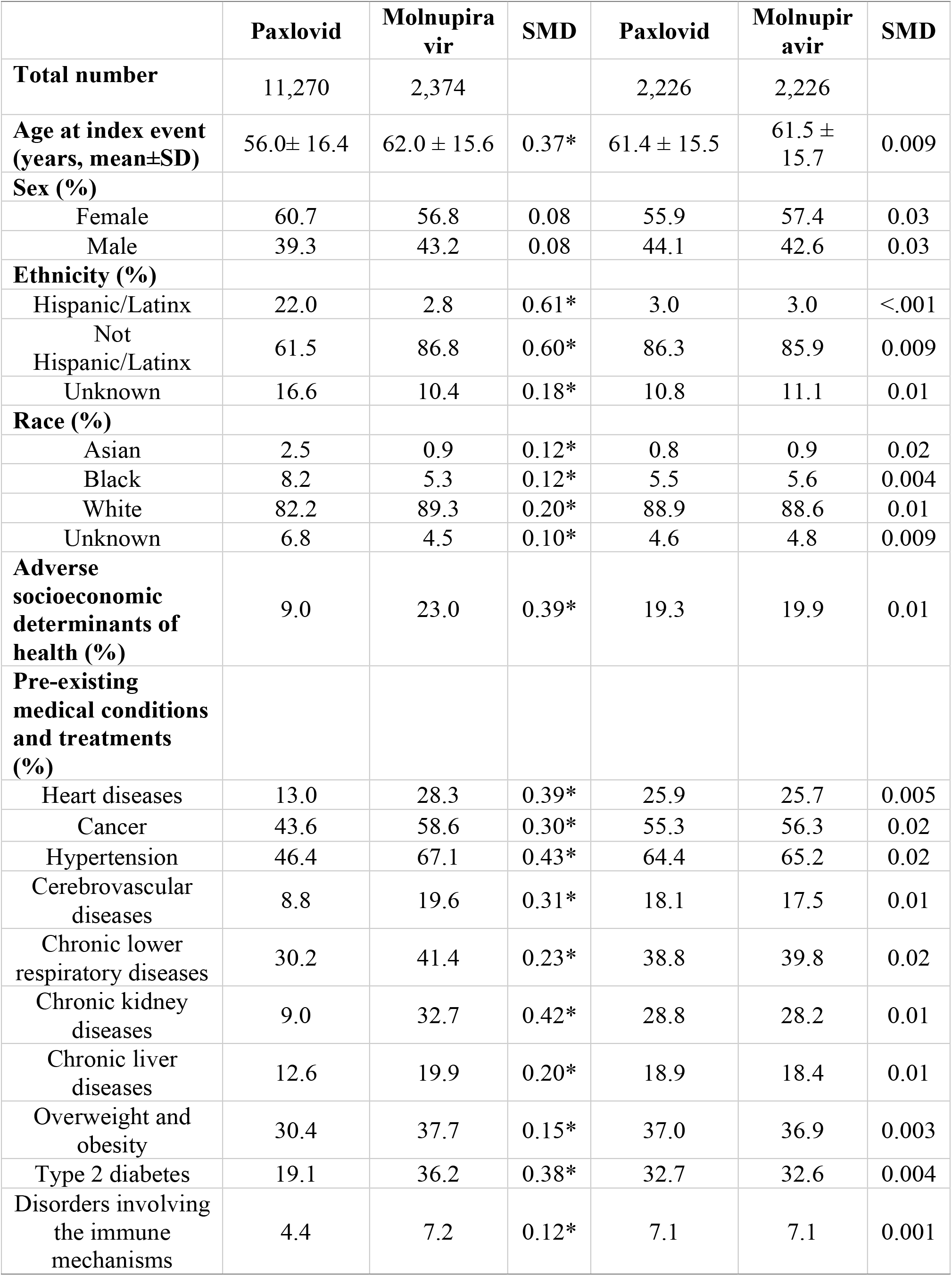
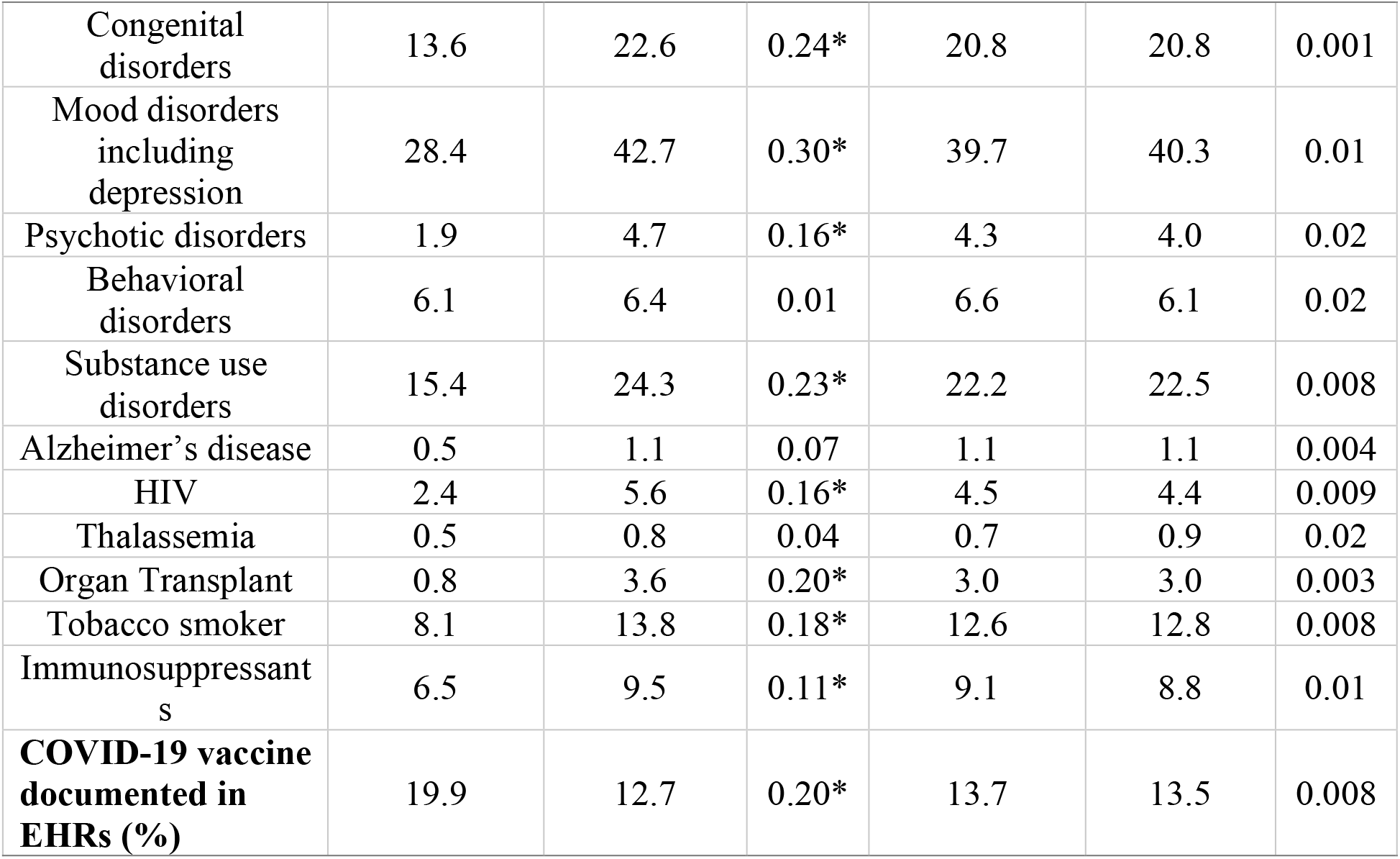
Characteristics of patients (aged ≥ 18 years old) before and after propensity-score matching (1:1 matching based on greedy nearest-neighbour matching with a caliper of 0.25 x standard deviation). Paxlovid – patients who contracted COVID-19 anytime between 1/1/2022- 6/8/2022 and were treated with Paxlovid within 5 days of COVID-19 diagnosis. Molnupiravir – patients who contracted COVID-19 anytime between 1/1/2022-6/8/2022 and were treated with Molnupiravir within 5 days of COVID-19 diagnosis. The status for adverse socioeconomic determinants of health, medical conditions, medications, procedures, and EHR-based COVID-19 vaccination status were based on presences of related codes in patient EHRs anytime up to 1 day before Paxlovid or Molnupiravir treatment. SMD – standardized mean differences. *SMD greater than 0.1, a threshold being recommended for declaring imbalance.

1. We compared risks for COVID-19 rebound outcomes in patients who took Paxlovid (“Paxlovid cohort”) and in patients who took Molnupiravir (“Molnupiravir cohort”). Three rebound outcomes (COVID-19 infections, COVID-19 related symptoms, hospitalizations) were followed for 7 days (from 2 through 8 days after the last day of treatment) and for 30 days (from 2 through 31 days after the last day of treatment) in the Paxlovid and the Molnupiravir cohorts. Kaplan-Meier analysis was used to estimate COVID-19 rebound outcomes. Cox’s proportional hazards model was used to compare the two matched cohorts with the proportional hazard assumption being tested with the generalized Schoenfeld approach. Hazard ratio (HR) and 95% confidence intervals was used to describe the relative hazard of rebound outcomes based on comparison of time to event rates. Separate analysis was performed in the two cohorts before and after propensity-score matching. For propensity-score matching, the two cohorts were 1:1 matched using a nearest neighbor greedy algorithm with a caliper of 0.25 times the standard deviation for variates described above.
2. We compared characteristics of patients who experienced COVID-19 rebound to those who did not experience rebound within 30 days (from 2 through 31 days after the last day of the medications) for both Paxlovid and Molnupiravir. Characteristics for comparison included demographics, adverse socioeconomic determinants of health, medical conditions, immunosuppressant usage, organ transplant procedures, and EHR-based COVID-19 vaccination status. All statistical tests were conducted within the TriNetX Advanced Analytics Platform. The TriNetX platform calculates HRs and associated CIs using R’s Survival package, version 3.2- 3. P-values are generated from a Z-test for present/absent variables, a t-test for continuous variables, and a Z-test for each category of categorical variables using the Python library SciPy.

## Results

### Patient characteristics

The study population comprised 13,644 patients aged ≥ 18 years old who contracted COVID-19 anytime between 1/1/2022-6/8/2022 (Omicron predominance period) and took Paxlovid (n = 11,270) or Molnupiravir (n=2,374) within 5 days of COVID-19 diagnosis. Patients treated with Paxlovid were younger than those treated with Molnupiravir (average age 56.0 vs 62.0). These two cohorts differed in gender, race, ethnicity, adverse socioeconomic determinants of health, pre-existing medical conditions including comorbidities, immunosuppressant usages, organ transplants and tobacco smoking, and EHR-documented COVID-19 vaccination status. After propensity-score matching, the two cohorts were balanced (Table 1).

### Rates and relative risks for COVID-19 rebound after Paxlovid vs Molnupiravir

Among 11,270 patients treated with Paxlovid, 398 (3.53%) tested positive, 260 (2.31%) had COVID-19 related symptoms and 50 (0.44%) were hospitalized during the 7-day period of from 2 through 8 days after the last day of Paxlovid. COVID-19 rebound rates were higher in the 2,374 patients treated with Molnupiravir: 5.86% for rebound infections, 3.75% for rebound symptoms and 0.84% for hospitalizations (**Figure 1a, top panel**). As shown in Table 1, patients who took molnupiravi were older and had more comorbidities. After matching, 7-day risks for COVID-19 rebound in patients treated with Paxlovid did not differ from those treated with Molnupiravir: rebound infections (HR: 0.81, 95% CI: 0.63-1.05), rebound symptoms (HR: 0.74, 95% CI: 0.53-1.03), and hospitalizations (HR: 0.78, 95% CI: 0.40-1.51) (**Figure 1b, bottom panel**).

**Figure 1.**
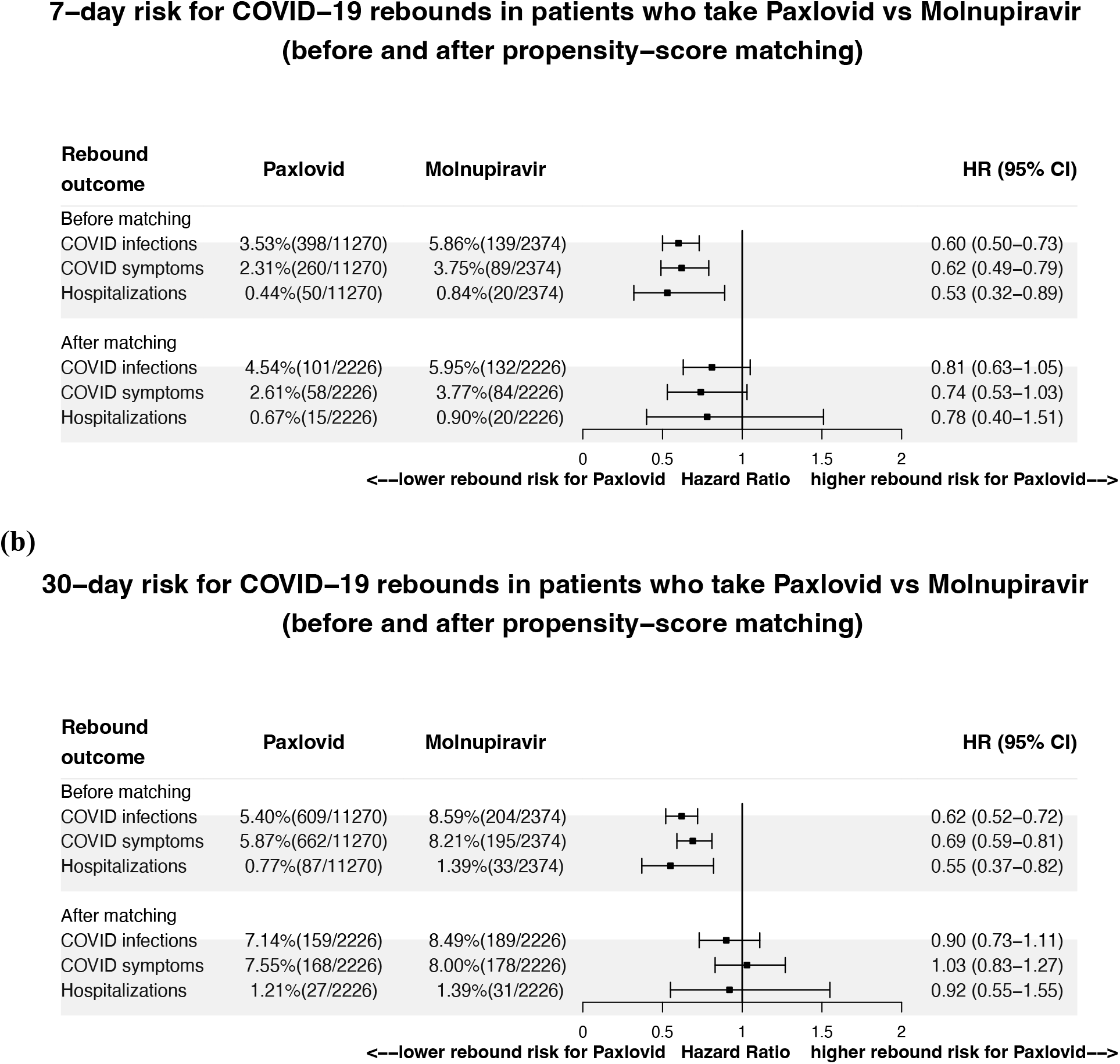
Comparison of 7- and 30-day risks for COVID-19 rebound in patients treated with Paxlovid or with Molnupiravir before and after propensity-score matching for demographics, adverse socioeconomic determinants of health, comorbidities, immunosuppressant usage, organ transplantation, and EHR-documented COVID-19 vaccination status. Rebound outcomes (COVID-19 infections, COVID-19 related symptoms, and hospitalizations) were followed for 7 days (from 2 through 8 days after the last day of treatments) and for 30 days (from 2 through 31 days after the last day of treatments).

For both drugs, the rebound rates increased as the time elapsed from the time of treatment. During the 30-day period of from 2 through 31 days after the last day of Paxlovid, 609 (5.40%) tested positive, 662 (5.87%) had COVID-19 related symptoms and 87 (0.77%) were hospitalized. For Molnupiravir, 30-day COVID-19 rebound rates also increased and they were higher than for Paxlovid: 8.59% for rebound infections, 8.21% for rebound symptoms and 1.39% for hospitalizations (**Figure 1b, top panel**). However, after matching, the 30-day risks for COVID- 19 rebound did not differ between the two drug treatments (**Figure 1b, bottom panel)**.

In summary, COVID-19 rebound (infection, symptoms, and hospitalizations) occurred for both drug treatments. The risks for rebound did not differ between Paxlovid and Molnupiravir, indicating that COVID-19 rebound is not unique to Paxlovid. The rebound rates increased as the time elapsed after treatment, suggesting inadequate viral clearance by the treatments.

### Characteristics of patients who had COVID-19 rebound vs those who did not

We compared characteristics of patients who developed COVID-19 rebound to those who did not within 30 days (from 2 through 31 days after the last day of treatment) for both Paxlovid and Molnupiravir. For Paxlovid, patients who developed COVID-19 rebound did not differ in age, or race from those without rebound, but there were more women and fewer Hispanics, had significantly more comorbidities, organ transplants and immunosuppressant usage and there were more tobacco smokers. The EHR-documented COVID-19 vaccination rate was higher in patients with COVID-19 rebound than those without, suggesting that vaccination is not a major contributor to COVID-19 rebound (**Table 2**). Similar trends were observed for Molnupiravir.

**Table 2.** Characteristics of patients with and without COVID-19 rebound. Age was based on current age as of June 19, 2022. The status for adverse socioeconomic determinants of health, medical conditions that are related to COVID-19 infection and outcomes including comorbidities, immunosuppressant usage, transplants, tobacco smoking, and COVID-19 vaccination status recorded in patient electronic health records were based on presences of related codes in patient EHRs anytime up to June 19, 2022.

|  | Paxlovid |  |  | Molnupiravir |  |  |
| --- | --- | --- | --- | --- | --- | --- |
|  | Rebound | No rebound | P-value | Rebound | No rebound | P-value |
| **Total number** | 609 | 10,662 |  | 204 | 2,170 |  |
| **Current age (as of 6/19/2022) (years, mean±SD)** | 57.9± 16.4 | 56.1 ± 16.4 | 0.07 | 63.6 ± 15.9 | 62.1 ± 15.5 | 0.10 |
| **Sex (%)** |  |  |  |  |  |  |
| Female | 65.2 | 60.5 | 0.02 | 57.8 | 56.7 | 0.76 |
| Male | 34.8 | 39.5 | 0.02 | 42.2 | 43.3 | 0.76 |
| **Ethnicity (%)** |  |  |  |  |  |  |
| Hispanic/Latinx | 16.7 | 22.2 | 0.001 | 4.9 | 2.8 | 0.08 |
| Not Hispanic/Latinx | 72.4 | 62.0 | <.001 | 90.7 | 86.5 | 0.09 |
| Unknown | 10.8 | 16.9 | <.001 | 6.4 | 10.8 | 0.05 |
| **Race (%)** |  |  |  |  |  |  |
| Asian | 1.3 | 2.5 | 0.17 | 0.5 | 0.9 | 0.81 |
| Black | 9.0 | 8.1 | 0.43 | 5.9 | 5.2 | 0.68 |
| White | 81.8 | 82.2 | 0.77 | 87.7 | 89.4 | 0.47 |
| Unknown | 7.6 | 6.8 | 0.46 | 5.9 | 4.3 | 0.31 |
| **Adverse socioeconomic determinants of health (%)** | 15.3 | 8.8 | <.001 | 27.5 | 22.7 | 0.12 |
| **Pre-existing medical conditions (%)** |  |  |  |  |  |  |
| Heart diseases | 20.5 | 12.8 | <.001 | 32.8 | 28.3 | 0.17 |
| Cancer | 54.8 | 43.3 | <.001 | 62.7 | 58.5 | 0.24 |
| Hypertension | 57.6 | 46.4 | <.001 | 73.0 | 67.1 | 0.09 |
| Cerebrovascular diseases | 16.7 | 8.5 | <.001 | 24.5 | 19.4 | 0.08 |
| Chronic lower respiratory diseases | 43.0 | 30.4 | <.001 | 54.4 | 41.2 | <.001 |
| Chronic kidney diseases | 15.4 | 8.9 | <.001 | 39.7 | 32.3 | 0.01 |
| Chronic liver diseases | 22.3 | 12.4 | <.001 | 28.4 | 19.4 | 0.002 |
| Overweight and obesity | 40.4 | 30.5 | <.001 | 43.6 | 37.9 | 0.11 |
| Type 2 diabetes | 30.0 | 18.8 | <.001 | 40.2 | 35.6 | 0.19 |
| Disorders involving the immune mechanisms | 9.0 | 4.6 | <.001 | 13.7 | 7.4 | 0.002 |
| Congenital disorders | 20.2 | 13.4 | <.001 | 25.0 | 22.5 | 0.41 |
| Mood disorders including depression | 40.1 | 28.0 | <.001 | 50.5 | 42.3 | 0.02 |
| Psychotic disorders | 3.3 | 1.8 | 0.009 | 6.9 | 4.6 | 0.15 |
| Behavioral disorders | 7.4 | 6.0 | 0.18 | 7.8 | 6.3 | 0.38 |
| Substance use disorders | 18.9 | 15.4 | 0.02 | 31.4 | 23.9 | 0.02 |
| Alzheimer's disease | 0.7 | 0.4 | 0.64 | 2.0 | 1.0 | 0.33 |
| HIV | 4.4 | 2.3 | <.001 | 5.4 | 5.6 | 0.89 |
| Thalassemia | 0.2 | 0.5 | 0.38 | 0.5 | 0.8 | 0.91 |
| Organ Transplant | 2.0 | 0.7 | <.001 | 4.9 | 3.5 | 0.41 |
| Tobacco smoker | 10.3 | 8.1 | 0.05 | 15.7 | 13.7 | 0.44 |
| Immunosuppressants | 10.3 | 6.4 | <.001 | 13.7 | 9.1 | 0.04 |
| **COVID-19 vaccine documented in EHRs (%)** | 24.8 | 19.6 | 0.002 | 17.6 | 12.2 | 0.03 |

While patients treated with Molnupiravir who developed rebound had more comorbidities than those without, the differences were not significant, which may reflect the small sample size of the Molnupiravir cohort. In addition, patients with rebound had higher EHR-documented vaccination rate than those without rebound.

In summary, patients with COVID-19 rebound had higher prevalence of comorbidities that are known to be associated with higher risk for COVID-19 infection and for adverse outcomes than those without rebound. This is consistent for both Paxlovid and Molnupiravir. There were no marked demographic differences between patients with and without rebound. Patients without rebound had higher prevalence of adverse socioeconomic determinants of health. For both drugs, patients with rebound had higher vaccination rates recorded in their EHR records.

## Discussion

Paxlovid and Molnupiravir were authorized by FDA in December 2021 to treat mild-to-moderate COVID-19 in patients who are at high risk for progression to severe COVID-19, with Paxlovid for 12 years or older and Molnupiravir for 18 years or older^1,2^. Our study population comprised patients age ≥ 18 years who contracted COVID-19 between 1/1/2022-6/8/2022 and were treated with Paxlovid or Molnupiravir within 5 days of COVID-19 infection. More people were prescribed Paxlovid (n = 11,270) than Molnupiravir (n=2,374), which may reflect the different outcomes between the two medications for cutting hospitalizations or death for high-risk patients as compared with placebo, corresponding to 88% for Paxlovid vs 30% for Molnupiravir^8^. Though both drugs were approved for infected patients at high risk for severe COVID-19, for our two cohorts the patients treated with Paxlovid differed significantly from those treated with Molnupiravir (Table 1). Patients treated with Paxlovid were significantly younger than those treated with Molnupiravir (average age 56.0 vs 62.0) and had fewer comorbidities. The Paxlovid cohort comprised more women, Hispanics, Asian and Black patients.

Our study shows that COVID-19 rebound was not unique to Paxlovid and occurred also in patients treated with Molnupiravir. The 30-day rebound rates were higher for Molnupiravir than Paxlovid: 8.59% vs 5.40% for rebound infections, 8.21% vs 5.87% for rebound symptoms and 1.39% vs 0.77% for hospitalizations. However, patients who took Molnupiravir were significantly older and had more comorbidities than those who took Paxlovid. After propensity-score matching, there were no significant differences in COVID-19 rebound risks between the two treatment cohorts. These results further suggest that rebound was not unique to Paxlovid and may be associated with persistent viral infection in some patients treated with either of these two antivirals. There has been more attention to COVID-19 rebounds following Paxlovid treatment than Molnupiravir^3,4^, which may be attributable to more people being treated with Paxlovid. Before propensity-score matching, patients who took Molnupiravir had higher hospitalization risks than those who took Paxlovid (1.39% vs 0.77%) during the time period of from 2 through 31 days after the last day of treatment, which is consistent with the reported higher trial efficacy results for Paxlovid (88%) than for Molnupiravir (30%)^8^. However, in the reported trials, both drugs were compared to placebo. Our results that compared Paxlovid to Molnupiravir in propensity-score matched patients showed no significant differences in either the 7-day or the 30-day risks for hospitalizations after treatments.

The rates of COVID-19 rebound for both drugs increased with time after treatments. For Paxlovid, the rate of COVID-19 infection rebound increased from 3.53% for 7 days to 5.40% for 30 days, a 53% increase. Similarly for Molnupiravir COVID-19 infection rebound rate increased from 5.86% for 7 days to 8.59% for 30 days, an 46.6% increase. This increase could occur if patients had inadequate viral clearance after treatment, patients did not complete the prescribed course of treatment or developed adverse drug effects and terminated treatment, if the dose was insufficient given pharmacodynamics in that individual, if reinfection occurred, or if viruses developed resistance to the drug. Future research is required to determine if this is the case and to evaluate instances when longer treatment duration might be indicated

Our study shows that patients with COVID-19 rebound were similar in age as those without rebound, but had significantly more comorbidities, organ transplants and immunosuppressant usage and more use of tobacco, suggesting that high risk patients with underlying medical conditions are more vulnerable to COVID-19 rebound. Future work is needed to dissect how each medical condition, for example, cancer, heart disease or type 2 diabetes, contributes to COVID-19 rebound while controlling for other factors. In the early phase of pandemic, studies showed that Black or African Americans and Hispanics were disproportionately impacted by COVID-19^9–14^. However, we observed no such racial and ethnic differences in COVID-19 rebound, suggesting COVID-19 treatments and vaccinations narrowed or eliminated the racial and ethnic gaps in COVID-19 infections and severe outcomes.

The rate of vaccination documented in patient EHRs were low (Table 1) compared to the actual vaccination rate of 89.5% in population ≥ 18 years of age^15^. This low recorded vaccination rate might be partially attributable to the fact that most vaccinations were performed outside of healthcare organizations and so were not recorded in the EHRs. Nonetheless, after propensity-score matching, the vaccination rates were balanced in the Paxlovid and the Molnupiravir cohorts. Given the high actual vaccination rate and balanced vaccination rates in propensity-score matched cohorts, the limited vaccination status captured in patient EHRs should not substantially impact our overall findings and conclusions. Our study showed that the vaccination rates were higher in patients who developed COVID-19 rebound than in those who did not, suggesting that vaccination was not a major contributor for COVID-19 rebound. While both drugs were tested in clinical trials that included only un-vaccinated populations, our study provided evidence that rebound occurred in largely vaccinated (89.5%) real-world populations and that rebound increased over time.

Since both drugs were approved for patients who are at high risk for COVID-19, it is not surprising that there is high prevalence of conditions associated with increased risk for COVID- 19 infection and severe outcomes (Table 1). For example, 43.6% and 46.4% patients treated with Paxlovid had cancer or hypertension respectively. The rates were even higher in patients treated with Molnupiravir: 58.6% had cancer and 67.1% had hypertension. It is unknown how the rate at which COVID-19 rebound develop in other, less compromised populations. However, COVID- 19 rebound occurred disproportionately in patients with pre-existing medical conditions (Table 2). The risk-benefit analysis of Paxlovid and Molnupiravir treatments in different populations warrants further investigation in real-world population by considering benefits of the drugs in preventing hospitalizations and deaths, and in developing COVID-19 rebound and drug resistance over time. This study population comprised patients who contracted COVID-19 anytime between 1/1/2022-6/8/2022, an Omicron predominant period. As the virus continues to evolve, we need to closely monitor how rebound develop in patients who are infected with different virus variants in the future. Due to sample size limitation, we did not differentiate among COVID-19 infected patients between those who were first time infected and those who were re-infected. As more people are getting reinfected, it will be important to examine whether COVID-19 rebound differ in reinfected and first-time infected patients.

Our study has several limitations: First, this is a retrospective observational study, so no causal inferences can be drawn. Second, there are inherent limitations in studies based on patient EHRs including over/mis/under-diagnosis and unmeasured confounders such as compliance with medication adherence and completion of treatment regimes. However, we compared the risks for COVID-19 rebound between the two cohort populations were both drawn from the TriNetX dataset, therefore these issues should not substantially affect the relative risk analyses. Third, patients in the TriNetX database represented those who had medical encounters with healthcare systems contributing to the TriNetX Platform. Even though this platform includes 28% of US population, it does not necessarily represent the entire US population. Therefore, results from the TriNetX platform need to be validated in other populations.

In summary, COVID-19 rebound occurred in patients treated with Paxlovid or with Molnupiravir, especially in those with underlying medical conditions. COVID-19 rebound is not unique to Paxlovid and the risks were similar for Paxlovid and Molnupiravir. The rates of COVID-19 rebounds increased with time after the treatments. Studies are necessary to determine the mechanisms underlying COVID-19 rebounds and to test dosing and duration regimes that might prevent such rebounds in vulnerable patients.

## Data Availability

All data produced in the present work are contained in the manuscript

## Contributors

RX conceived and designed the study and drafted the manuscript. LW performed data analysis and prepared tables and figures and participated in manuscript preparation. NDV, NAB, PBD, DCK critically contributed to study design, result interpretation and manuscript preparation. We confirm the originality of content. RX had full access to all the data in the study and takes responsibility for the integrity of the data and the accuracy of the data analysis.

## Acknowledgments

We acknowledge support from National Institute on Aging (grants nos. AG057557, AG061388, AG062272, AG076649), National Institute on Alcohol Abuse and Alcoholism (grant no. R01AA029831), the Clinical and Translational Science Collaborative (CTSC) of Cleveland (grant no. 1UL1TR002548-01), National Cancer Institute Case Comprehensive Cancer Center (R25CA221718, P30 CA043703, P20 CA2332216).

## Meeting Presentation

No

